# Genomic Analysis and Surveillance of Respiratory Syncytial Virus (RSV) Using Wastewater-Based Epidemiology (WBE)

**DOI:** 10.1101/2023.07.21.23293016

**Authors:** Danielle M. Allen, Marina I. Reyne, Pearce Allingham, Ashley Levickas, Stephen H. Bell, Jonathan Lock, Jonathon D. Coey, Stephen Carson, Andrew J. Lee, Cormac McSparron, Behnam Firoozi Nejad, James McKenna, Mark Shannon, Kathy Li, Tanya Curran, Lindsay J. Broadbent, Damian G. Downey, Ultan F. Power, Helen E. Groves, Jennifer M. McKinley, John W. McGrath, Connor G. G. Bamford, Deirdre F. Gilpin

## Abstract

Respiratory syncytial virus (RSV) causes severe infections in infants, immunocompromised or elderly individuals resulting in annual epidemics of respiratory disease. Currently, limited clinical RSV surveillance and the lack of predictable RSV seasonal dynamics and limits the public health response. Wastewater-based epidemiology (WBE) has the capacity to determine levels of health-associated biomarkers and has recently been used globally as a key metric in determining prevalence of SARS-CoV-2 in the community. However, the application of genomic WBE for the surveillance of other respiratory viruses is limited.

In this study, we present an integrated genomic WBE approach, using RT-qPCR and partial sequencing of the G gene to monitor RSV levels and variants in the community across 2 years encompassing two periods of high RSV clinical positivity in Northern Ireland.

We report increasing detection of RSV in wastewater concomitant with increasing numbers of RSV positive clinical cases. Furthermore, analysis of wastewater-derived RSV sequences permitted subtyping, genotyping, and identification of distinct circulating lineages within and between seasons.

Altogether, our genomic WBE platform has the potential to complement ongoing global surveillance efforts and aid the management of RSV by informing the timely deployment of pharmaceutical and non-pharmaceutical interventions.

## Introduction

Respiratory syncytial virus (RSV), an enveloped, non-segmented, negative-sense RNA virus, is a major cause of respiratory illness in humans, placing substantial burden on healthcare systems worldwide (1). Globally, RSV is estimated to cause ∼3.1 million hospitalisations annually, which, in 2015, resulted in ∼118,000 deaths in children under the age of 5 and ∼14,000 deaths in adults ≥65 years (2,3). Disease management in the United Kingdom (UK) is currently limited to prevention, symptomatic relief and short-term protection for high-risk infants by administration of monthly immunoprophylaxis (1). However, the introduction of nirsevimab, a long-acting monoclonal antibody, and several promising vaccine candidates, for older adults and pregnant women, could protect individuals from RSV-related hospitalisation during the RSV season (4,5).

RSV causes seasonal epidemics, typically occurring between October and March in the northern hemisphere (6). Clinical disease surveillance usually relies on the identification of infections in individuals with severe symptoms and/or co-morbidities where hospitalisation is required (7). During the coronavirus disease (COVID-19) pandemic, RSV seasonal occurrence changed dramatically with little or no evidence of RSV circulation in the first year of the pandemic, followed by a surge in off-season outbreaks across the globe (8), presumably due to imposed non-pharmaceutical interventions in response to the global pandemic (9). Limited clinical RSV surveillance and lack of predictable RSV seasonal dynamics and limits the public health response, impacting clinical resource planning and allocation.

Furthermore, compared to other pathogens, like SARS-CoV-2 and Influenza A virus (IAV), genomic RSV surveillance is either lacking or typically focuses on paediatric clinical samples (10,11). With the expected introduction of new immunoprophylaxis and vaccines, continued surveillance of RSV genotypes circulating within the community could help identify antigenic variations, potential immune escape mutations, and track the effectiveness of available prophylactics (including the emergence of potential resistance) (4,8). Therefore, the establishment of a robust surveillance system to rapidly determine the onset of the RSV ‘season’, the genotypes of circulating variants and both its community spread and the prevalence of RSV between seasons could support public health decision making regarding the initiation of immunoprophylaxis at the onset of the season.

Wastewater-based epidemiology (WBE) gained popularity as a public health surveillance system during the COVID-19 pandemic by supporting governmental policies and promoting quicker actions to prevent and control the spread of SARS-CoV-2 (12,13). Many studies have shown that wastewater surveillance can identify the magnitude, distribution, and community-level prevalence of SARS-CoV-2, irrespective of whether individuals are symptomatic or not (12,14). Indeed, detection and quantification of SARS-CoV-2 RNA concentrations significantly correlated with clinical cases and hospitalisations within the same wastewater catchment area, while sequence analysis facilitated the tracking of variants and identification of cryptic lineages (15,16).

Beohm *et al.,* (2022) and Hughes *et al.,* (2022) have shown that RSV can be detected in wastewater settled solids, with the latter also demonstrating that RNA concentrations in settled solids correlated with clinical trends in RSV positivity rates (17,18). More recently, Ahmed *et al.,* (2023) reported correlations between wastewater RSV concentrations and clincal cases (19). We now describe the implementation of a WBE approach for RSV surveillance, based in Northern Ireland (NI), to track the community spread of RSV during two successive seasons (2021 and 2022). As with previous studies, RSV levels in wastewater correlated to RSV positive clinical cases. Using a sequence-based approach and phylogenetics, we compared wastewater-derived RSV A and B G gene sequences and linked clinical sequences to infer transmission dynamics within and between seasons.

## Methods

### Sample collection, viral concentration, extraction, and RT-qPCR

Between August 2021–February 2023, composite samples of primary untreated influent were collected weekly from twenty wastewater treatment works (WWTW) across NI, covering 57% of the population (Figure S1, Table S1). An Isco Glacier autosampler (Lincoln, USA) was used by NI Water Ltd to collect 200 mL of sample every 15 min over a 24 h period before delivery to Queen’s University Belfast by the NI Environment Agency. For RSV detection, wastewater samples were concentrated and the nucleic acid was extracted as previously described (20). RT-qPCR was performed on the QuantStudio**™** 7 Real-Time PCR System (ThermoFisher Scientific) in triplicate using an RSV A and B N gene assay described previously (18). A sample was considered positive if one out of three replicates amplified. A standard curve and limit of detection (LOD) were calculated for the assay (Protocol 1, Figure S2). RSV RNA concentrations were normalised using wastewater flow rate and expressed as gene copies (g.c.) per 100,000 population equivalents (p.e.) per day (20).

### Statistical analysis

To reduce variability in the measurements a B-spline model was used to smooth the RSV g.c. per 100,000 p.e. per day (21). Different degrees of splines and sequence of knot sets were tested to smooth the data, and the final model was selected based on the Bayesian Information Criterion (BIC). The number of positive RSV clinical cases per week for NI between August 2021–February 2023 were provided by the Regional Virology Lab (RVL), Belfast Health and Social Care Trust (BHSCT). The relationship between RSV concentration in wastewater and clinical cases was investigated using linear regression. All analyses and data visualization were performed in R v4.2.2. (R Core Team 2021) unless otherwise stated.

### Amplicon sequencing of RSV G gene

Positive wastewater samples and 6 clinical samples from October 2022 (provided by the RVL) were sequenced. Extracted nucleic acid was reverse transcribed with LunaScript RT SuperMix Kit (New England BioLabs). The thermocycling conditions were 25°C for 2 min, 55°C for 20 min, 95°C for 1 min before cooling to 4°C. The cDNA was amplified in two subsequent PCR reactions (external and semi-nested) targeting the second half of the hypervariable G gene (Table S2) (22). The reaction and thermocycling conditions used are shown in Table S3-4.

Amplicons were analysed using the TapeStation D1000 assay (Agilent, CA) and samples with the appropriate fragment size (Table S2) were taken forward for sequencing. Libraries were prepared following the minimized Nextera XT library preparation protocol (23) and sequenced on an Illumina MiSeq instrument using a MiSeq v2 300 cycles reagent kit. Reads were pre-processed by removing adaptors and low-quality reads (below Q30). Reads representing each sample were mapped onto the RSV A isolate 0594 (GenBank MW582528.1) and RSV B isolate 9671 (GenBank MW582529.1) to generate consensus sequences, which are recent dominant circulating genotypes harbouring characteristic G gene subregion duplication (24). Finally, the sequences were deposited on GISAID (Accession numbers: XXX).

### Sequence analysis

Multiple sequence alignments using partial G gene sequences were performed in MAFFT [https://mafft.cbrc.jp/] (25), and maximum likelihood phylogenies inferred (1000 bootstrap replicates) using IQtree [http://iqtree.cibiv.univie.ac.at/] (26). The best substitution model was identified using IQtree, ranked by BIC (GTR+F+G4 (A) and TIM3+F+I+G4 (B)). High coverage RSV A and B genome sequences collected between 2017 and 2022, were downloaded from GISAID database (7th November 2022), matching G gene sequences extracted using BLAST and analysed alongside the sequences generated in this study (27). Trees were visualised using FigTree v1.4.4 [http://tree.bio.ed.ac.uk/software/figtree/]. Table S5 provides metadata on the GISAID genome sequences used in this study. Genotype predictions were carried out using Nextclade online tool (https://clades.nextstrain.org/) (28). Relevant aligned nucleic acid sequences were translated into the predicted protein sequence and visualised using Jalview (29).

## Results

Two RSV epidemics were captured in NI (August 2021–January 2022, and July 2022–February 2023, respectively) (Fig. 1A). During the 2021 season, RSV RNA was first detected in August 2021 at an average concentration of 1.31×10^10^ g.c. per 100,000 p.e. per day across the 20 WWTW. The RSV concentration subsequently increased and peaked in September 2021 at an average concentration of 4.93×10^11^ g.c. per 100,000 p.e. per day. The RSV concentration in wastewater decreased from October 2021 and was no longer detected by January 2022. RSV was detected sporadically in wastewater samples between the 2021 and 2022 RSV seasons (Fig. 1B). During the 2022 season, RSV concentration increased and peaked in October 2022 at an average concentration of 1.25×10^12^ g.c. per 100,000 p.e. per day. Across this sampling period, the first peak in RSV positive clinical cases was observed in September 2021 and the second peak was observed in October 2022. This correlated with the temporal pattern of RSV gene fragment detection within wastewater: a strong relationship was observed between RSV RNA concentration in wastewater and PCR positive cases from infected individuals (R^2^ = 0.730). The geographic distribution of RSV RNA across the 20 WWTW in NI varied in magnitude and spread within and between each season (Fig. 1B). For example, in 2021 RSV RNA concentrations were highest in County Armagh (WWTW: Lurgan, Craigavon and Armagh) compared to 2022 where the RSV RNA concentrations were highest in County Tyrone, Londonderry, and Antrim (WWTW: Omagh, Limavady and North Coast).

**Figure 1.**
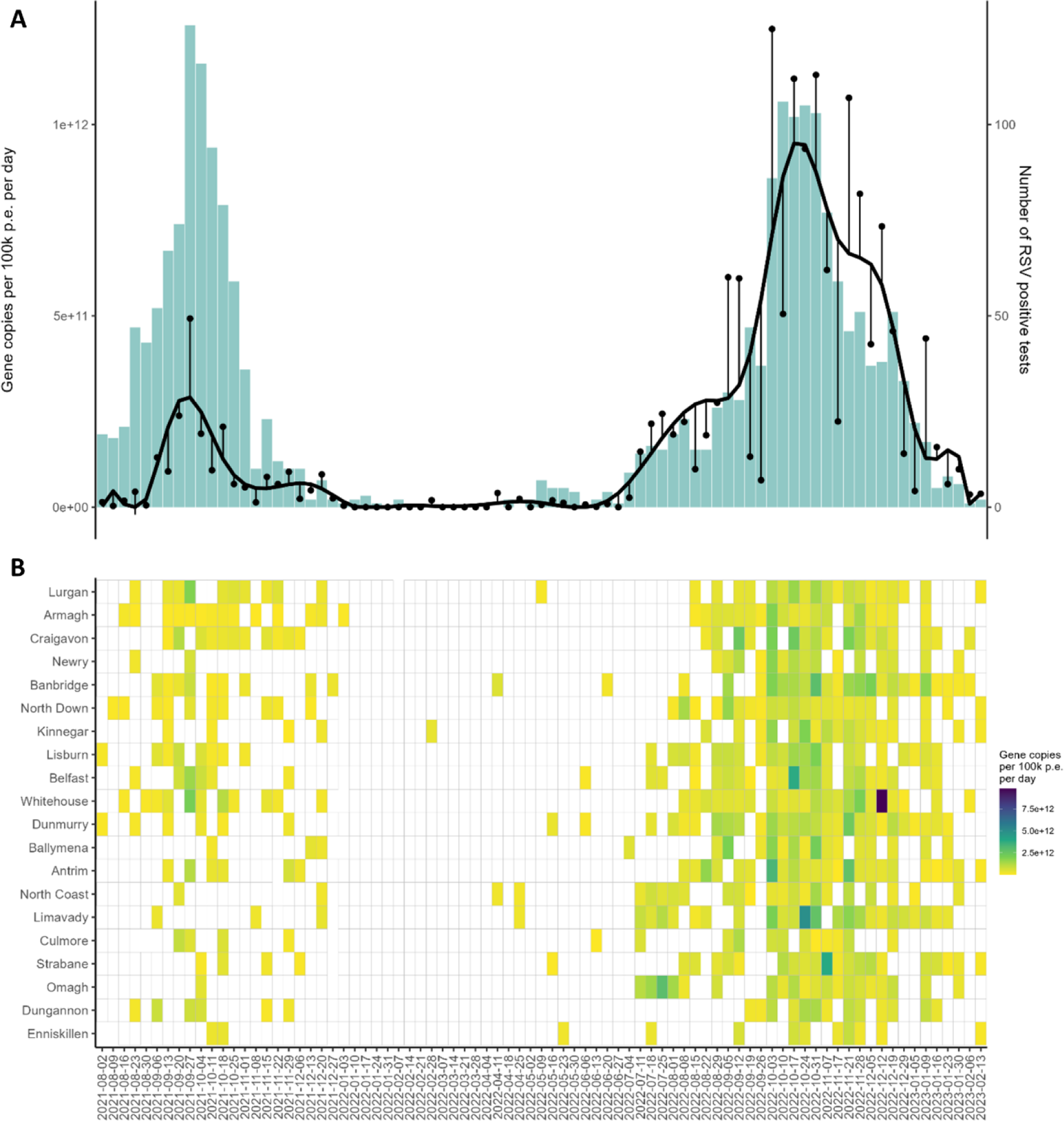
A) Averaged RSV RNA concentrations (g.c. per 100,000 p.e. per day) across 20 WWTW in NI (black circles) and smoothed data based on fitting a B spline model (black line) from August 2021– February 2023. Histogram showing the number of RSV positive cases for NI during the same period. B) Heat map showing the distribution of RSV RNA gene copies per 100,000 p.e. per day in wastewater across the 20 WWTW in NI between August 2021–February 2023.

RSV can be subdivided into two subtypes and numerous genotypes, and differences have been noted between subtypes (30). Our RT-qPCR assay targets a conserved portion of the N gene (18); therefore, to provide greater resolution of RSV diversity and evolution, the second hypervariable region of the G gene (corresponding to mucin-like domain II) was sequenced from positive wastewater samples (22). This region of G is an established genetic region used in RSV surveillance (31). Overall, 10 RSV A and 14 RSV B consensus sequences were derived from wastewater and 2 RSV A and 4 RSV B consensus sequences were derived from clinical samples. Further analysis of the sequencing process, the sequencing depth for each sample, and further details on the wastewater-derived RSV A and B consensus sequences are detailed in Tables S6-S8.

RSV A was detected more frequently in the 2021 season, whereas RSV B was detected more frequently in the 2022 season. The predicted genotypes were clade A3 (G clade GA2.3.5) for all RSV-A sequences, and B6 (G clade GB5.0.5a) for all RSV-B sequences. A more detailed phylogenetic analysis of derived sequences, in the context of contemporary global RSV sequences, suggested the presence of several distinct local clades of both genotypes (A3 and B6) across both seasons in NI. For both RSV A and B, 3 major local clades were detected, herein referred to as RSV A1-3 and B1-3 (Figure 2). Clades A1 and B3 were further subdivided into two groups each (X.1 and X.2) due to intra-clade genetic differences. Clinical sequences were found in the same clade as WW sequences. In the 2021 season, all 3 distinct clades of RSV A were found from wastewater samples (dominated by A1) and only 1 RSV B clade (B3.1) while during the 2022 season, only 1 RSV A clade (A1.2) and all 3 RSV B clades (B1, 2 and B3.2) were found (dominated by B1). NI RSV sequences often clustered with other contemporary sequences detected globally but, in some instances, sequences from both seasons clustered only with NI sequences (A1.2 and B3). Clade A1 is subdivided into A1.1 and A1.2 sister lineages with A1.1 being large and diverse containing NI sequences from both 2021 and 2022, and sequences found outside NI between 2019-2022. In contrast, A1.2 formed its own cluster of mostly NI sequences from both 2021 and 2022, which were found in sister lineages, and a single Australian sequence from 2019 that sits basal to all A1.2 NI sequences. A2 contains two NI sequences on a long branch with no clear close relatives in other clades. However, the most closely related sequences were from sequences isolated in Argentina and Philippines from 2019. A3 contains a single NI sequence closely related to a sequence from Egypt from 2019. Like A1, B1 is a large and diverse clade containing NI sequences alongside global sequences from 2021 and 2022. Interestingly, UK sequences from 2019 and 2020 sit basal with respect to the rest of the clade. Clade B2 contains a single NI sequence as a sister lineage to recent Australian sequences. Clade B3 is composed only of NI sequences but interestingly from both 2021 and 2022 and is split into B3.1 (2021) and B3.2 (2022) by the presence of a long branch linking the seasons.

**Figure 2:**
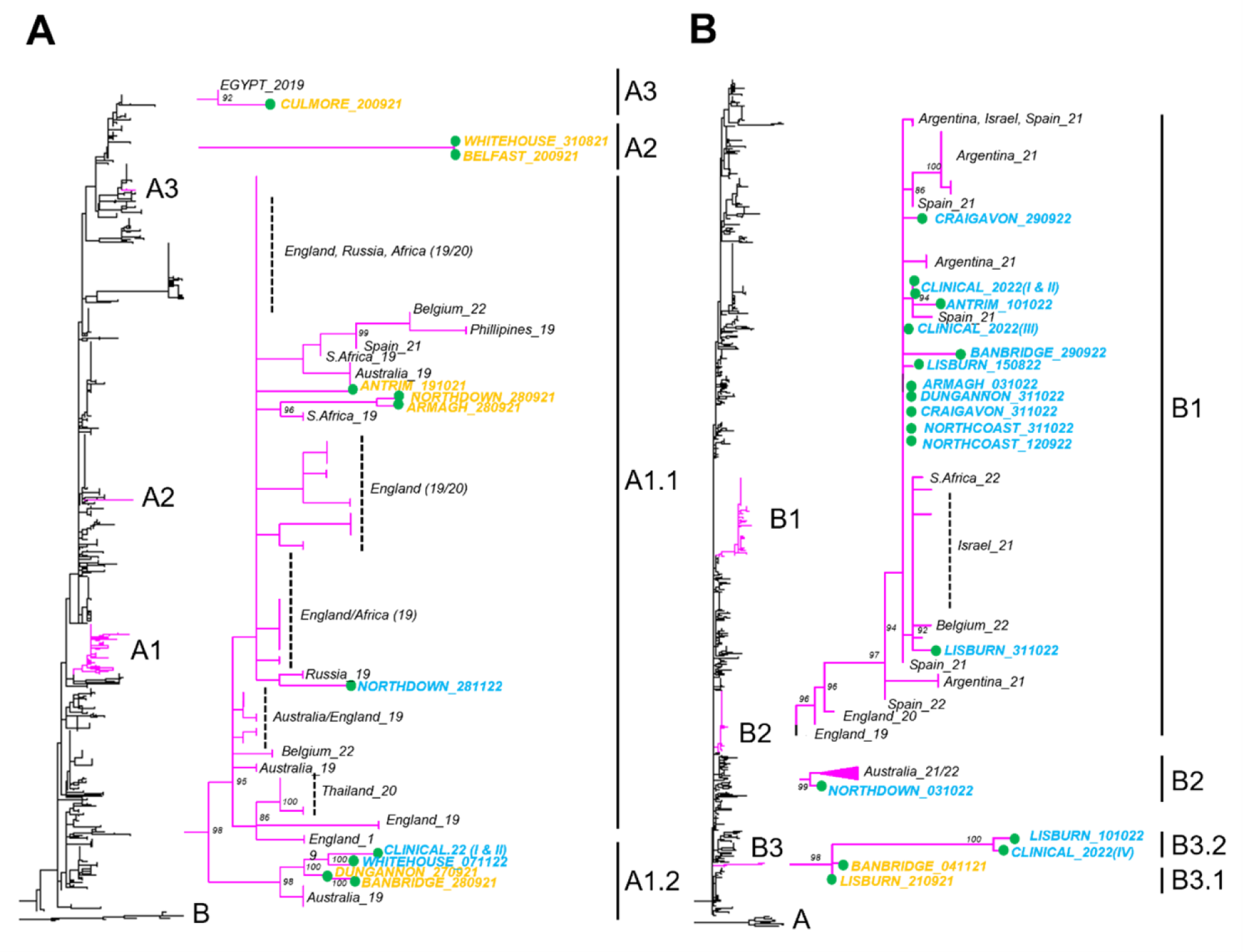
Maximum likelihood phylogenetic trees of generated NI RSV A (A) and B (B) based on G gene sequences alongside contemporary sequences (available through GISAID), with 1000 bootstrap replicates using IQtree. Whole phylogeny shown on left with sequences generated in this study highlighted (pink) alongside focused clades containing NI sequences (inset). In inset, taxa are labelled with date and geographical origin metadata for all derived NI sequences and for certain global sequences. For clarity, bootstraps over 70 are shown only.

Analysis of the predicted encoded amino acid sequences for our sequences, and other select non-NI sequences within our clusters reinforced the presence of at least 3 clades per subtype (Fig. 3). While differences were noted across the region, focusing between seasons within a clade we identified several amino acids changes, like S251F in A1.2. In B3.2 there were 9 amino acid mutations (L219P, L252P, D253N, I254T, L286P, Y287H, I290T, STOP313Q). Considering the emergence of clade B1 after 2020, we identified several derived mutations preceding its emergence (I254T, K258N, I270T, S277P, Y287H, R314K). Finally, we identified several amino acid changes shared between B clades including: L219P (B1 and B3.2) and STOP313Q (B1 and B3.2), and most prominently in I254T (B1 and B3.2) and Y287H (B1 and B3.2). Furthermore, mutations at 253 and 258 could result in the addition of an extra N-linked glycosylation site in both B1 and B3.2 in a similar region. Additionally, several mutations are noted that are likely to affect O-linked glycosylation patterns through gain (I254T in B1 and B3.2, I270T in B1, I290T in B1) or loss (S251F in A1.2, T267P in B3.2, and S277P in B1) of sites. No amino acid differences specific to clinical sequences were identified compared to related wastewater sequences.

**Figure 3:**
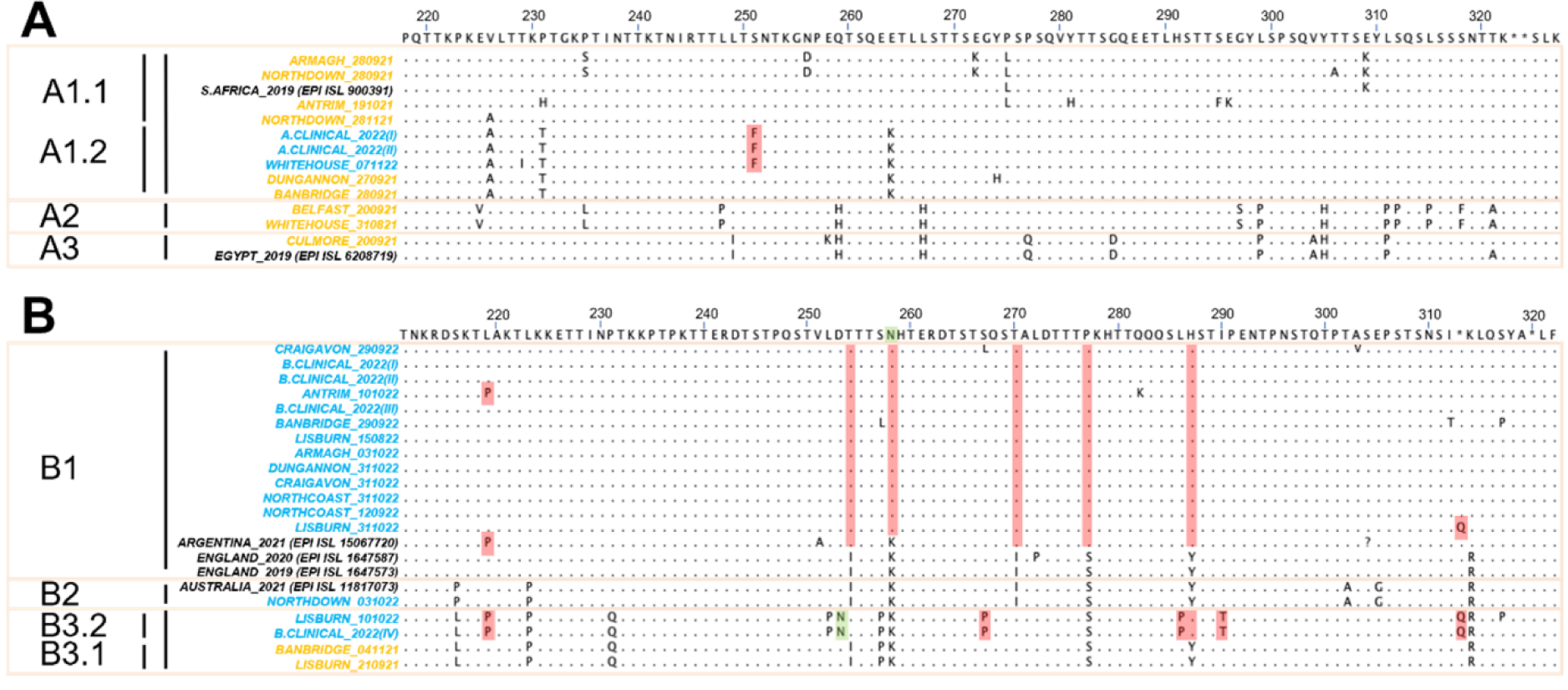
Encoded amino acid sequences for RSV A (A) and RSV B (B) samples from this study and other relevant non-NI sequences within our clusters were visualised in JalView. Amino acid sequences were separated into groups based on the clades from the phylogenetic analysis. Mutations shown in reference to the consensus of all sequences (41). Amino acids the same as consensus were highlighted with a dot. Stop codons were shown (*). Mutations of interest were shaded in red. N-linked glycosylation site were shaded in green.

Integrating our WBE platform, the wastewater-derived RSV sequences from NI were mapped across time and space (Fig. 4). This revealed that the RSV lineages were not evenly dispersed across NI, with most lineages detected in County Down, Armagh and Antrim. Interestingly, the greatest diversity in lineages was detected in County Down, with 7 different RSV lineages found over the 2021 and 2022 season. While the dominant lineages like A1 and B1 were widespread, other lineages were only found in 1 county, including A3 in September 2021, B2 and B3.2 in October 2022. To visualise RSV genetic diversity over time, and the relationship to viral load in wastewater, a dashboard was created: http://go.qub.ac.uk/RSV-NI

**Figure 4:**
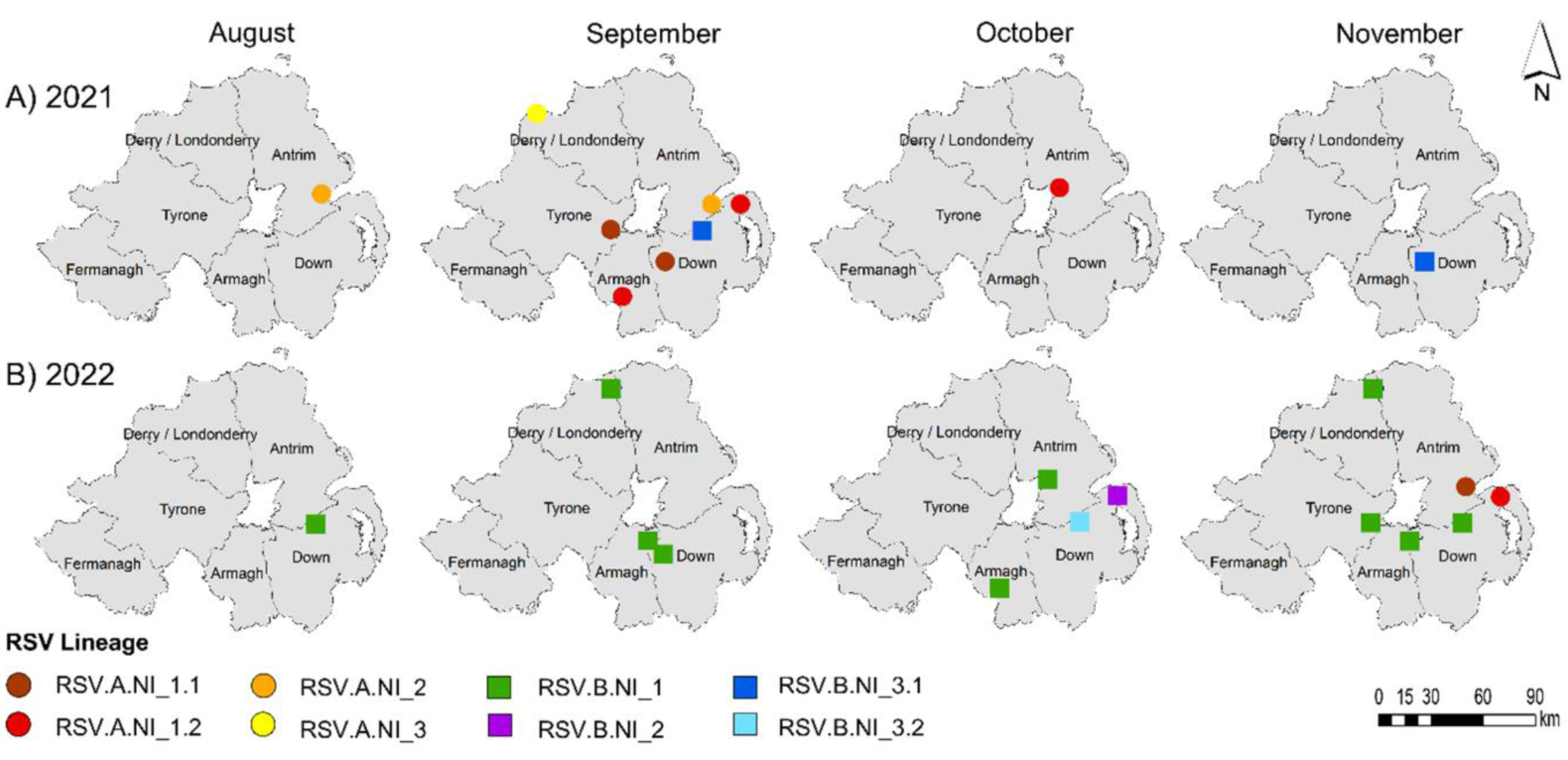
The geospatial detection of the RSV A and B lineages across 2021 and 2022 seasons in NI, divided into six counties (Antrim, Armagh, Fermanagh, Tyrone, Londonderry and Down). RSV A and B lineages were shown as circles or squares. The different RSV A and B lineages (A1.1, A1.2, A2, A3, B1, B2, B3.1 and B3.2) were shown in different colours and shades.

## Discussion

This study demonstrated that composite influent wastewater samples followed by population normalisation can be used to successfully monitor the prevalence of RSV in wastewater. The RSV RNA concentration in wastewater correlated with RSV positive clinical cases. Furthermore, this study captured two unusual RSV epidemics between August 2021–February 2023, which were inconsistent with the traditional winter seasonal patterns observed in the UK (32). This was consistent with off-season RSV epidemics reported worldwide following the removal of COVID-19 pandemic restrictions (8). Importantly, the data generated from RSV wastewater surveillance could aid the RSV clinical response by determining the onset of the RSV ‘season’, thereby supporting public health campaigns to raise awareness regarding prevention of RSV infection and transmission. Furthermore, these data could help inform clinical decision making regarding the initiation of palivizumab immunoprophylaxis of high risk infants at the onset of the RSV ‘season’. In general, higher RSV RNA concentrations were observed in 2022 compared to 2021, while the number of reported RSV positive clinical cases were similar. Several factors could have contributed to these differences, including clinical testing only detecting severe disease and therefore, the burden of RSV infections in the community infected is unknown. It is unknown if RSV shedding rates in faeces differ between ages, disease state, or RSV subtype, and if these factors differed between epidemics then RSV RNA concentrations in the wastewater would vary.

This study highlights the utility of WBE for tracking RSV genetic diversity, as evidenced by the epidemiologically-relevant RSV sequences derived from wastewater targeting the G gene. RSV seasons in NI were characterised by the introduction and circulation of multiple lineages. This included both subtypes (RSV A and B), represented by a single genotype each but encompassing several genetically distinct clades, consistent with other studies (31,33). In this study, the NI sequences were consistent with sequences that were concomitantly circulating worldwide, and hence reflective of global circulation patterns. The spread of acute respiratory infections is strongly influenced by the global movement of people between the southern and northern hemispheres, as well as the tropics (31). Thus, it is hard to precisely determine the origin of NI RSV strains, but speculatively, it is likely that much of the observed RSV genetic diversity within NI was derived from outside the country.

We further sub-divided our sequences into 3 local clades per genotype/subtype. This is likely an underestimate given that within some clades we find phylogenetic structuring suggestive of additional importation events from outside NI (e.g. within clade A1.1). One interesting observation of our study was the identification of sequences within clades A1.2 and B3 where the most closely related sequences between epidemics were derived from within NI. This pattern is consistent with epidemiological persistence of RSV that has been noted previously (34). Indeed, we detected RSV in wastewater samples between seasons, but it was sporadic both spatially and temporally. However, the factors that contribute to the re-emergence of RSV in the community are still poorly understood, with ambient temperature, humidity and sunlight thought to play a role on stability and infectivity of RSV, while human behaviour, strain variation, and the introduction of novel strains is also believed to be integral to the transmission (35,36). Interestingly, within A1.2, the 2021 and 2022 sequences were in sister clades, which might suggest re-introduction from an unknown source, potentially even outside of NI. Additionally, the link between B3.1 and B3.2 is on a long branch, with multiple mutations between them, rendering interpretations regarding the origin of the strain difficult.

Importantly, this study highlighted that wastewater-derived RSV sequences were found in similar clades as RSV sequences derived from clinical samples. As such, wastewater sequencing accurately reflects the genotypes circulating in the entire community and causing clinical disease. Interestingly, other studies have found that SARS-CoV-2 wastewater-derived sequence data indicated that more lineages were circulating across the sampled communities than were represented in the clinical-derived data (16,37). Therefore, expanding RSV genomic surveillance to encompass both clinical and environmental samples will greatly expand our understanding of the genetic and epidemiological characteristics of the virus at the population level, while also rapidly identifying antigenic variations and potential immune escape mutations. This ability to follow RSV circulation and evolution at the population level will undoubtedly become even more important following the imminent introduction and widespread use of long-acting prophylactic monoclonal antibodies and RSV vaccines (38).

While RSV G gene sequencing can provide clues into the origin and spread of RSV within NI, it can also provide insights into virus biology as although the highly glycosylated hyper-variable domain (mucin-like region II) of G is largely unstructured, the G protein is involved in binding and entry of RSV (39). Through this, we identified several findings of note, of which the apparent convergent evolution in RSV B was the most striking. Several variable N-glycosylation sites were identified within B1 and B3.2, which potentially could affect G biology and potential immune evasion (38,40).

There were several limitations to this study. First, pre-pandemic samples were not available for inclusion. Second, only ∼10% of wastewater samples were successfully sequenced using the protocol developed in this study. The reasons for this are unclear, with wastewater being a complex sample type containing PCR inhibitors, and variation in viral load within wastewater potentially contributing. Third, while the wastewater-derived sequences closely matched the clinical-derived sequences, only small numbers of clinical samples were obtained from the 2022 season and none for the 2021 season. Fourth, the geographical location and date of clinical test data was unavailable and therefore, it was not possible to compare spatially with the wastewater lineages. Fifth, the positivity rate for the clinical samples was also unavailable. Finally, the phylogenetic analysis in this study was based on the second hypervariable region of the G gene. However, F gene or whole genome sequencing would provide further information on the genetic and epidemiological characteristics of the virus. Therefore, future work could include a large longitudinal wastewater surveillance study to assess the antigenic evolution of RSV between and within seasons over many years.

Overall, this study highlights the value of WBE as an effective tool to detect and monitor the onset and circulation of RSV in the community. Its implementation will undoubtedly be of considerable use to public health teams worldwide to make rapid pharmaceutical and non-pharmaceutical interventions to help control RSV outbreaks. Furthermore, continued surveillance of co-circulating RSV genotypes using WBE could help identify antigenic variations and potential immune escape mutations against future pharmaceutical interventions, including long-acting monoclonal antibodies, vaccines and antivirals.

## Data Availability

All data produced in the present study are available upon reasonable request to the authors

## Acknowledgements

We gratefully acknowledge all data contributors, i.e., the Authors and their Originating laboratories responsible for obtaining the specimens, and their Submitting laboratories for generating the genetic sequence and metadata and sharing via the GISAID Initiative, on which this research is based. We are thankful to Northern Ireland (NI) Water Ltd and RPS Group for wastewater sample collection, and the Department of Agriculture, Environment and Rural Affairs, NI Environment Agency, Department for Infrastructure and Department of Finance.

## Ethical statement

This study was conducted using environmental samples and publicly available data. Under UK law, there is no requirement for ethical approval for these samples or data.

## Data availability

N/A

## Corresponding author

Danielle Allen: School of Biological Sciences, Queen’s University Belfast, 97 Lisburn Road, Belfast, BT9 7BL, Northern Ireland (UK).

