## Supplementary material for "Genomic Analysis and Surveillance of Respiratory Syncytial Virus (RSV) Using Wastewater-Based Epidemiology (WBE)"

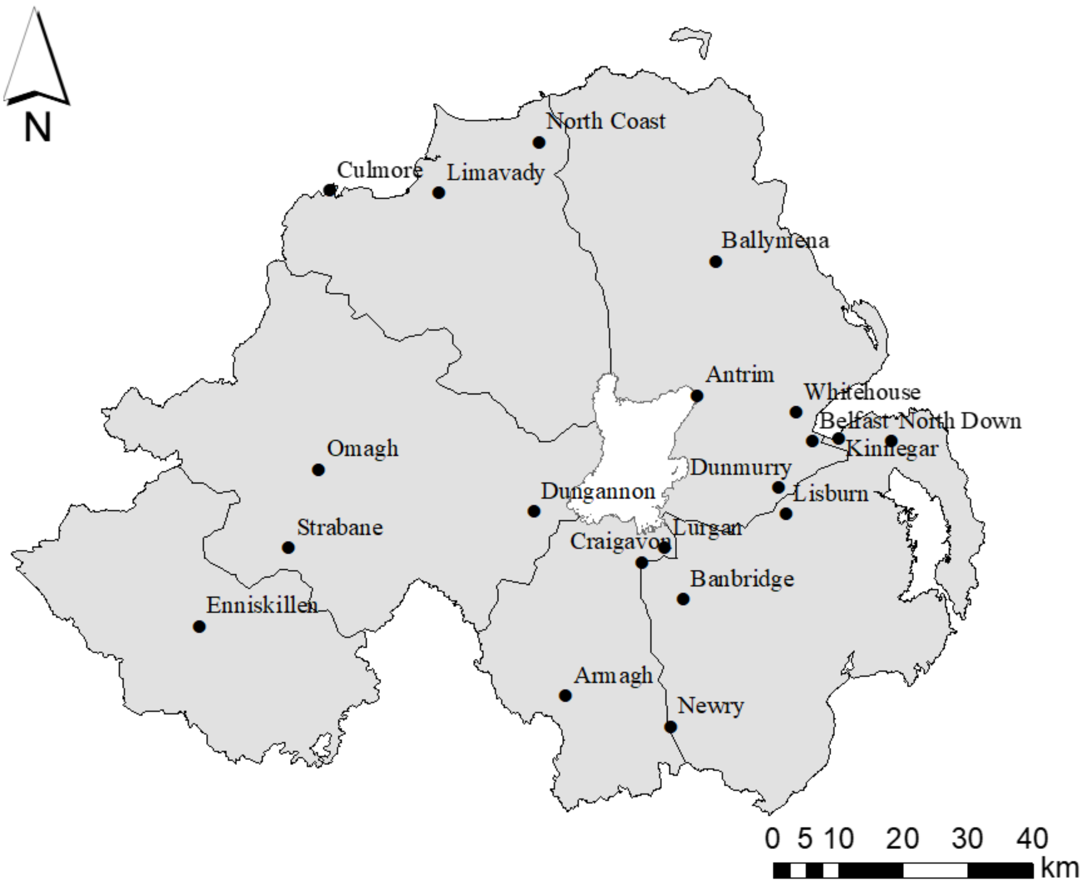


**Figure S1.** Map showing geographical locations of sampled WWTW across Northern Ireland (NI).

**Table S1.** County and population equivalents (p.e.) for each WWTW

<table>
<tr><th>County</th><th>WWTW</th><th>Population equivalents (p.e.)</th></tr>
<tr><td rowspan="3">Londonderry</td><td>Culmore</td><td>94,655</td></tr>
<tr><td>Limavady</td><td>11,583</td></tr>
<tr><td rowspan="2">North Coast</td><td rowspan="2">42,440</td></tr>
<tr><td rowspan="6">Antrim</td></tr>
<tr><td>Ballymena</td><td>42,969</td></tr>
<tr><td>Antrim</td><td>41,735</td></tr>
<tr><td>Dunmurry</td><td>42,397</td></tr>
<tr><td>Whitehouse</td><td>68,925</td></tr>
<tr><td>Belfast</td><td>228,939</td></tr>
<tr><td rowspan="5">Down</td><td>Lisburn</td><td>47,377</td></tr>
<tr><td>Banbridge</td><td>18,803</td></tr>
<tr><td>North Down</td><td>73,384</td></tr>
<tr><td>Kinnegar</td><td>97,582</td></tr>
<tr><td>Craigavon</td><td>78,899</td></tr>
<tr><td rowspan="3">Armagh</td><td>Newry</td><td>34,042</td></tr>
<tr><td>Armagh</td><td>15,749</td></tr>
<tr><td>Lurgan</td><td>29,302</td></tr>
<tr><td rowspan="3">Tyrone</td><td>Omagh</td><td>20,200</td></tr>
<tr><td>Strabane</td><td>13,251</td></tr>
<tr><td>Dungannon</td><td>18,079</td></tr>
<tr><td>Fermanagh</td><td>Enniskillen</td><td>15,115</td></tr>
</table>

**Protocol 1.** Standard curve

RSV Clinical BT2a isolate was kindly supplied by Dr Lindsay Broadbent, isolation and characterisation of this isolate was previously described [1]. RSV B positive control was purchased from ZeptoMetrix (NATtrol RSV positive Control NATRSV-6C, ZeptoMetrix). RSV A and B were quantified using digital droplet PCR (ddPCR). For this assay, ddPCR was performed on 20 µl samples from a 22 µl reaction volume, prepared using a 5.5 µl template, mixed with 5.5 µl of One-Step RT-ddPCR Advanced Kit for Probes (Bio-Rad 1863021), 2.2 µl of 200 U/µl Reverse Transcriptase, 1.1 µl of 300 mM DTT, and primers and probes at a final concentration of 900 and 250 nM, respectively. Droplets were generated using the AutoDG Automated Droplet Generator (Bio-Rad). PCR was performed using C1000 Touch Thermal Cycler with the following cycling conditions: 50°C for 60 mins, 95°C for 10 mins, 40 cycles at 95°C for 30 sec and 55°C for 1 min, and 98 °C for 10 mins. Droplets were analysed using the QX200 Droplet Reader (Bio-Rad), and each well had to have over 10,000 droplets for inclusion in the analysis. Thresholding was done using the QX Manager Software (Bio-Rad, version 1.2). A standard curve was generated for RSV A and B N gene using sixfold serial dilutions of a quantified positive control.

1. Villenave R, O'Donoghue D, Thavagnanam S, Touzelet O, Skibinski G, Heaney LG, McKaigue JP, Coyle P V., Shields MD, Power UF. Differential cytopathogenesis of respiratory syncytial virus prototypic and clinical isolates in primary pediatric bronchial epithelial cells. *Virol. J.* BioMed Central Ltd; 2011; 8.

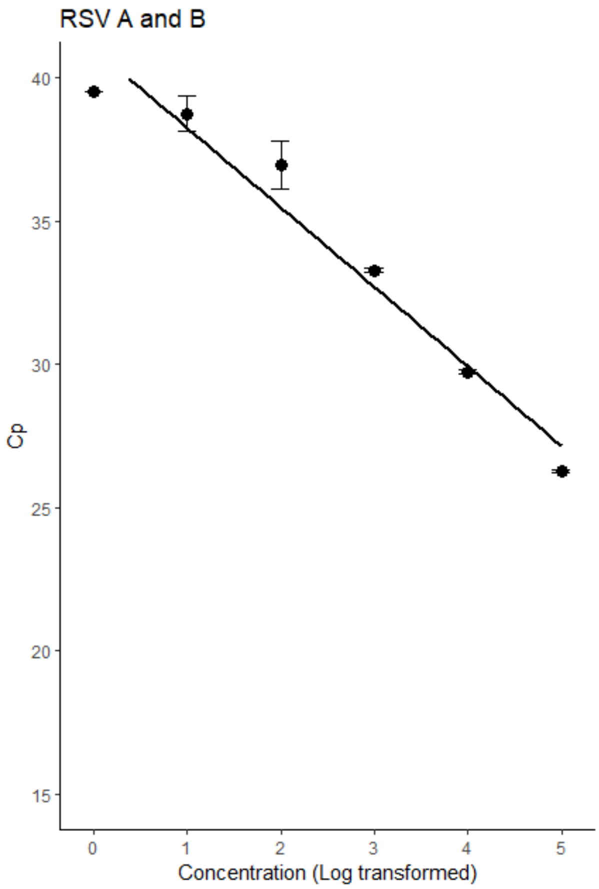


**Figure S2.** Standard curve for RSV A and B N gene. Amplification efficiency was 129%. The LOD for RSV RNA detection in wastewater was 1 copy per reaction

**Table S2.** Description of PCR primers used for the genotyping of RSV A and B in wastewater samples.

| Assay | Region | Primer name | Sequence (5' - 3') | Final reaction concn (nM) | Amplicon size (bp) |
|---|---|---|---|---|---|
| Sequencing: External PCR | G gene | ABG490 - Forward | ATGATTWYCAYTTTGAAGTGTTC | 500 | A: 607<br>B: 661<br>BA: 670 |
| | | F164 - Reverse | GTTATGACACTGGTATACCAACC | 500 | |
| Sequencing: Seminested PCR | G gene | AG655 – RSV A Forward | GATCYCAAACCTCAAACCAC | 500 | A: 460<br>B: 585<br>BA: 645 |
| | | BG517 – RSV B Forward | TTYGTTCCCTGTAGTATATGTG | 500 | |
| | | F164 – RSV A and B Reverse | GTTATGACACTGGTATACCAACC | 500 | |

**Table S3:** Reaction mixture conditions for external and semi-nested PCRs

| Reaction Mixture | Final reaction concn (nM) | 1 reaction (µL) |
|---|---|---|
| Reaction buffer (New England BioLabs) | | 5 |
| Q5 High-Fidelity DNA Polymerase (New England BioLabs) | | 0.25 |
| dNTPs (New England BioLabs) | 10 | 0.5 |
| Q5 High GC Enhancer (New England BioLabs) | | 5 |
| Forward primer (Eurofins) | 500 | 1.25 |
| Reverse primer (Eurofins) | 500 | 1.25 |
| Nuclease Free Water | | 9.25 |
| Template | | 2.5 |
| Total | | 25 |

**Table S4:** Thermocycling conditions for external and semi-nested PCRs

| Thermocycling conditions | External PCR | | | Semi-nested PCR for RSV A | | | Semi-nested PCR for RSV B | | |
|---|---|---|---|---|---|---|---|---|---|
| | Temperature (°C) | Time (s) | Number of cycles | Temperature (°C) | Time (s) | Number of cycles | Temperature (°C) | Time (s) | Number of cycles |
| Initial denaturation | 98 | 60 | | 98 | 60 | | 98 | 60 | |
| Annealing | 98 | 10 | 35 | 98 | 10 | 30 | 98 | 10 | 30 |
| | 54 | 30 | | 62 | 30 | | 56 | 30 | |
| | 72 | 40 | | 72 | 40 | | 72 | 40 | |
| Final extension | 72 | 120 | | 72 | 120 | | 72 | 120 | |
| Hold | 4 | ∞ | | 4 | ∞ | | 4 | ∞ | |

**Table S6**: Analysis of the sequencing process

| | RSV A | RSV B |
|---|---|---|
| Number of RT-qPCR positive wastewater samples selected for external and semi-nested PCR | 281 | 233 |
| Number of samples with expected band from the tape station analysis | 159 | 101 |
| Total number of consensus sequences obtained | 10 | 14 |
| Average Ct value and standard deviation of wastewater samples selected for external and semi-nested PCR | 37.15 ± 0.89 | 37.18 ± 0.93 |
| Average Ct value and standard deviation of wastewater samples that were successfully sequenced | 37.23 ± 0.86 | 36.75 ± 0.76 |

**Table S7:** The mean read depth per base ± standard deviation for each consensus sequence.

| RSV Serotype | Sample ID | Mean read depth per base ± standard deviation |
|---|---|---|
| A | AMH280921 | 1217 ± 789 |
| | ATM191021 | 2831 ± 1417 |
| | BEL200921 | 3107 ± 1904 |
| | BNB280921 | 2504 ± 1699 |
| | CUL200921 | 862 ± 777 |
| | DNG270921 | 2172 ± 1090 |
| | NDN280921 | 1479 ± 1294 |
| | WTH310821 | 7292 ± 1696 |
| | NDN281122 | 1817 ± 962 |
| | WTH071122 | 7081 ± 1917 |
| | V22034499 | 4740 ± 3482 |
| | V22034533 | 3969 ± 3498 |
| B | BNB041121 | 1186 ± 496 |
| | LIS210921 | 6575 ± 2398 |
| | AMH031022 | 6111 ± 2398 |
| | ATM101022 | 5942 ± 2726 |
| | BNB290922 | 6820 ± 2283 |
| | CRG290922 | 6037 ± 2480 |
| | CRG311022 | 6867 ± 2215 |
| | DNG311022 | 6639 ± 2252 |
| | LIS101022 | 6535 ± 2336 |
| | LIS150822 | 5974 ± 2295 |
| | LIS311022 | 6767 ± 2243 |
| | NCT120922 | 5176 ± 2071 |
| | NCT311022 | 6840 ± 2229 |
| | NDN031022 | 5841 ± 2783 |
| | V22033750 | 6210 ± 2186 |
| | V22034570 | 5263 ± 2136 |
| | V22034901 | 1855 ± 819 |
| | V22035143 | 6927 ± 2209 |

**Table S8**: Details on the RSV A and B consensus sequences derived from wastewater and clinical samples.

| Sample ID | Sample type | Year | WWTW | RSV Subtype | NextClade G clade | Assigned Lineage in this study |
|---|---|---|---|---|---|---|
| AMH280921 | Wastewater | 2021 | Armagh | A | GA2.3.5 | RSV.A.NI_1.2 |
| ATM191021 | Wastewater | 2021 | Antrim | A | GA2.3.5 | RSV.A.NI_1.2 |
| BEL200921 | Wastewater | 2021 | Belfast | A | GA2.3.5 | RSV.A.NI_2 |
| BNB280921 | Wastewater | 2021 | Banbridge | A | GA2.3.5 | RSV.A.NI_1.1 |
| CUL200921 | Wastewater | 2021 | Culmore | A | GA2.3.5 | RSV.A.NI_3 |
| DNG270921 | Wastewater | 2021 | Dungannon | A | GA2.3.5 | RSV.A.NI_1.1 |
| NDN280921 | Wastewater | 2021 | North Down | A | GA2.3.5 | RSV.A.NI_1.2 |
| WTH310821 | Wastewater | 2021 | Whitehouse | A | GA2.3.5 | RSV.A.NI_2 |
| BNB041121 | Wastewater | 2021 | Banbridge | B | GB5.0.5a | RSV.B.NI_3.1 |
| LIS210921 | Wastewater | 2021 | Lisburn | B | GB5.0.5a | RSV.B.NI_3.1 |
| NDN281122 | Wastewater | 2022 | North Down | A | GA2.3.5 | RSV.A.NI_1.2 |
| WTH071122 | Wastewater | 2022 | Whitehouse | A | GA2.3.5 | RSV.A.NI_1.1 |
| AMH031022 | Wastewater | 2022 | Armagh | B | GB5.0.5a | RSV.B.NI_1 |
| ATM101022 | Wastewater | 2022 | Antrim | B | GB5.0.5a | RSV.B.NI_1 |
| BNB290922 | Wastewater | 2022 | Banbridge | B | GB5.0.5a | RSV.B.NI_1 |
| CRG290922 | Wastewater | 2022 | Craigavon | B | GB5.0.5a | RSV.B.NI_1 |
| CRG311022 | Wastewater | 2022 | Craigavon | B | GB5.0.5a | RSV.B.NI_1 |
| DNG311022 | Wastewater | 2022 | Dungannon | B | GB5.0.5a | RSV.B.NI_1 |
| LIS101022 | Wastewater | 2022 | Lisburn | B | GB5.0.5a | RSV.B.NI_3.2 |
| LIS150822 | Wastewater | 2022 | Lisburn | B | GB5.0.5a | RSV.B.NI_1 |
| LIS311022 | Wastewater | 2022 | Lisburn | B | GB5.0.5a | RSV.B.NI_1 |
| NCT120922 | Wastewater | 2022 | North Coast | B | GB5.0.5a | RSV.B.NI_1 |
| NCT311022 | Wastewater | 2022 | North Coast | B | GB5.0.5a | RSV.B.NI_1 |
| NDN031022 | Wastewater | 2022 | North Down | B | GB5.0.5a | RSV.B.NI_2 |
| V22034499 | Clinical | 2022 | | A | GA2.3.5 | RSV.A.NI_1.1 |
| V22034533 | Clinical | 2022 | | A | GA2.3.5 | RSV.A.NI_1.1 |
| V22033750 | Clinical | 2022 | | B | GB5.0.5a | RSV.B.NI_1 |
| V22034570 | Clinical | 2022 | | B | GB5.0.5a | RSV.B.NI_3.2 |
| V22034901 | Clinical | 2022 | | B | GB5.0.5a | RSV.B.NI_1 |
| V22035143 | Clinical | 2022 | | B | GB5.0.5a | RSV.B.NI_1 |

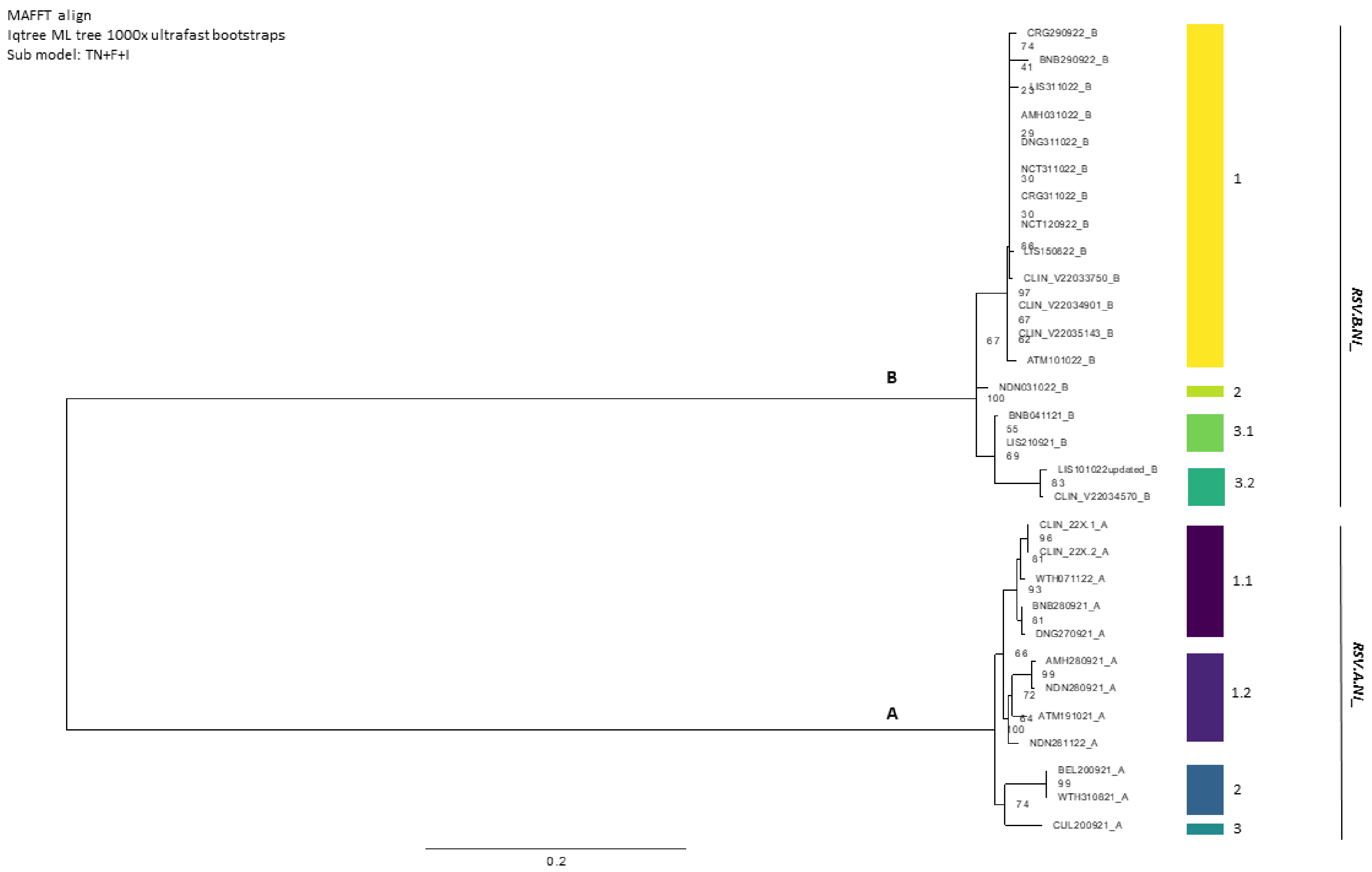


**Figure S3:** Maximum likelihood phylogenetic trees of generated NI RSV A and B based on G gene sequences with 1000 bootstrap replicates using IQtree.

**Figure S4 and Figure S5**: Maximum likelihood phylogenetic trees of generated NI RSV A (S4) and B (S5) based on G gene sequences alongside contemporary sequences (available through GISAID), with 1000 bootstrap replicates using IQtree. (PDF)
