## Supplementary figures and images for "Genomic Analysis and Surveillance of Respiratory Syncytial Virus (RSV) Using Wastewater-Based Epidemiology (WBE)"

### Supplementary Figure S4

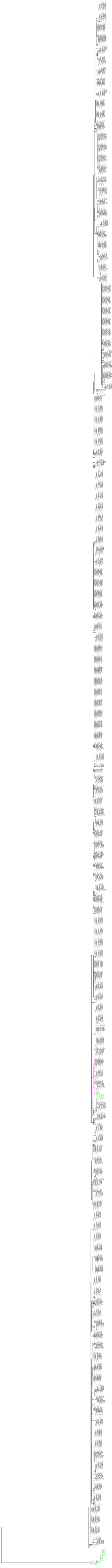

### Supplementary Figure S5

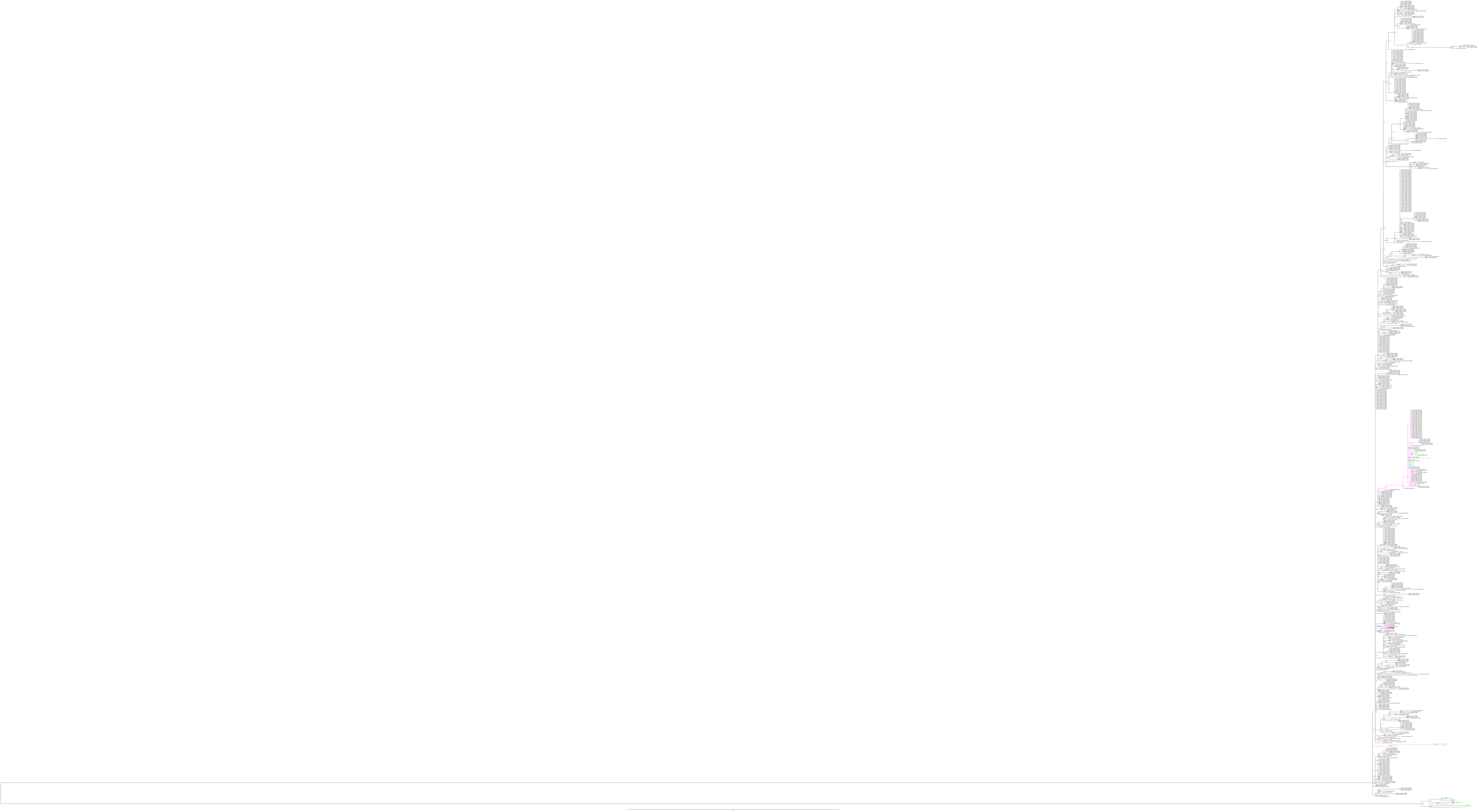
